# VinDr-PCXR: An open, large-scale chest radiograph dataset for interpretation of common thoracic diseases in children

**DOI:** 10.1101/2022.03.04.22271937

**Authors:** Ngoc H. Nguyen, Hieu H. Pham, Thanh T. Tran, Tuan N.M. Nguyen, Ha Q. Nguyen

## Abstract

Computer-aided diagnosis systems in adult chest radiography (CXR) have recently achieved great success thanks to the availability of large-scale, annotated datasets and the advent of high-performance supervised learning algorithms. However, the development of diagnostic models for detecting and diagnosing pediatric diseases in CXR scans is undertaken due to the lack of high-quality physician-annotated datasets. To overcome this challenge, we introduce and release VinDr-PCXR, a new pediatric CXR dataset of 9,125 studies retrospectively collected from a major pediatric hospital in Vietnam between 2020 and 2021. Each scan was manually annotated by a pediatric radiologist who has more than ten years of experience. The dataset was labeled for the presence of 36 critical findings and 15 diseases. In particular, each abnormal finding was identified via a rectangle bounding box on the image. To the best of our knowledge, this is the first and largest pediatric CXR dataset containing lesion-level annotations and image-level labels for the detection of multiple findings and diseases. For algorithm development, the dataset was divided into a training set of 7,728 and a test set of 1,397. To encourage new advances in pediatric CXR interpretation using data-driven approaches, we provide a detailed description of the VinDr-PCXR data sample and make the dataset publicly available on https://physionet.org/.

## Background & Summary

Common thoracic diseases cause several hundred thousand deaths every year among children under five years old^1,2^. The chest radiograph or CXR is the first-line and most commonly performed imaging examination in the assessment of the pediatric patient^3^. Interpreting CXR scans on pediatric patients can be for a number of indications or critical findings, in particular for common thoracic diseases in children such as Pneumonia, Bronchitis and Cardiovascular diseases (CVDs). Depending on the patients’ age, the difficulty of the examination will vary, often requiring a specialist in pediatric diagnostic imaging with an in-depth knowledge of radiological signs of different lung conditions^4^. Additionally, the inter-observer agreement and intra-observer agreement in the pediatric CXR interpretation were low^5^. This opens room for the development of data-driven approaches and computational tools to assist pediatricians in the diagnosis of common thoracic diseases and to reduce their workload.

Computer-aided diagnosis (CAD) systems for identification of lung abnormality in adult CXRs have recently achieved great success thanks to the availability of large labeled datasets^6–10^. Many large-scale CXR datasets of adult patients such as ChestX-ray14^6^, Padchest^7^, CheXpert^8^, MIMIC-CXR^9^ and VinDr-CXR^10^ have been established and released in recent years. These datasets boosted new advances in exploring new machine learning-based approaches in the interpretation of CXR in adults^8,11–16^. Unfortunately, the creation of pediatric CXR datasets is still unexploited, and the number of benchmark pediatric CXR datasets is limited. This becomes the main obstacle in developing and transferring new machine learning-based CAD systems for pediatric CXR in clinical practice.

In an effort to provide a large-scale pediatric CXR dataset with high-quality annotations for the research community, we have built the VinDr-PCXR dataset in DICOM format. The dataset consists of 9,125 posteroanterior (PA) view CXR scans in patients younger than 10 years that were retrospectively collected from three major hospitals in Vietnam from 2020 to 2021. In particular, all CXR scans come with both the localization of critical findings and the classification of common thoracic diseases. These images were annotated by a group of three radiologists with at least 10 years of experience for the presence of 36 critical findings (*local labels*) and 15 diagnoses (*global labels*). Here, the local labels should be annotated with rectangle bounding boxes that localize the findings, while the global labels reflect the diagnostic impression of the radiologist at the image-level. For algorithm development, we randomly divided the dataset into two parts: the training set of 7,728 scans (84.7%) and the test set of 1,397 scans (15.3%). To the best of our knowledge, the released VinDr-PCXR is currently the largest public pediatric CXR dataset with radiologist-generated annotations in both training and test sets. Table 1 below shows an overview of existing public datasets for CXR interpretation in pediatric patients, compared with the VinDr-PCXR. Compared to the previous works, the VinDr-PCXR dataset shows two main advantages. First, the dataset is labeled for multiple findings and diseases. Meanwhile, most pediatric CXR datasets have focused on a single disease such as pneumonia^17^ or pneumothorax^18^. Second, the dataset provides bounding box annotations at lesion level, which is useful for developing explainable artificial intelligent models^19^ for the CXR interpretation in children. We believe the introduction of the VinDr-PCXR provides a suitable imaging source for investigating the ability of supervised machine learning models in identifying common lung diseases in pediatric patients.

**Table 1.** An overview of existing public datasets for CXR interpretation in pediatric patients.

| Dataset | Release year | # findings | # samples | Image-level labels | Local labels |
| --- | --- | --- | --- | --- | --- |
| Kermany *et al.*<sup>17</sup> | 2018 | 2 | 5,856 | Available | Not available |
| Chen *et al.*<sup>18</sup> | 2020 | 5 | 2,668 | Available | Not available |
| VinDr-PCXR (ours) | 2021 | 52 | 9,125 | Available | Available |

## Methods

### Data collection

Data collection was conducted at the Phu Tho Obstetric & Pediatric Hospital (PTOPH) between 2020 – 2021. The ethical clearance of this study was approved by the Institutional Review Boards (IRBs) of the PTOPH. The need for obtaining informed patient consent was waived because this retrospective study did not impact clinical care or workflow at these two hospitals, and all patient-identifiable information in the data has been removed. We retrospectively collected more than 10,000 CXRs in DICOM format from a local picture archiving and communication system (PACS) at PTOPH. The imaging dataset was then transferred and analyzed at Smart Health Center, VinBigData JSC.

### Overview of approach

The building of the VinDr-PCXR dataset is illustrated in Figure 1. In particular, the collection and normalization of the dataset were divided into four main steps: (1) data collection, (2) data de-identification, (3) data filtering, and (4) data labeling. We describe each step in detail as below.

**Figure 1.**
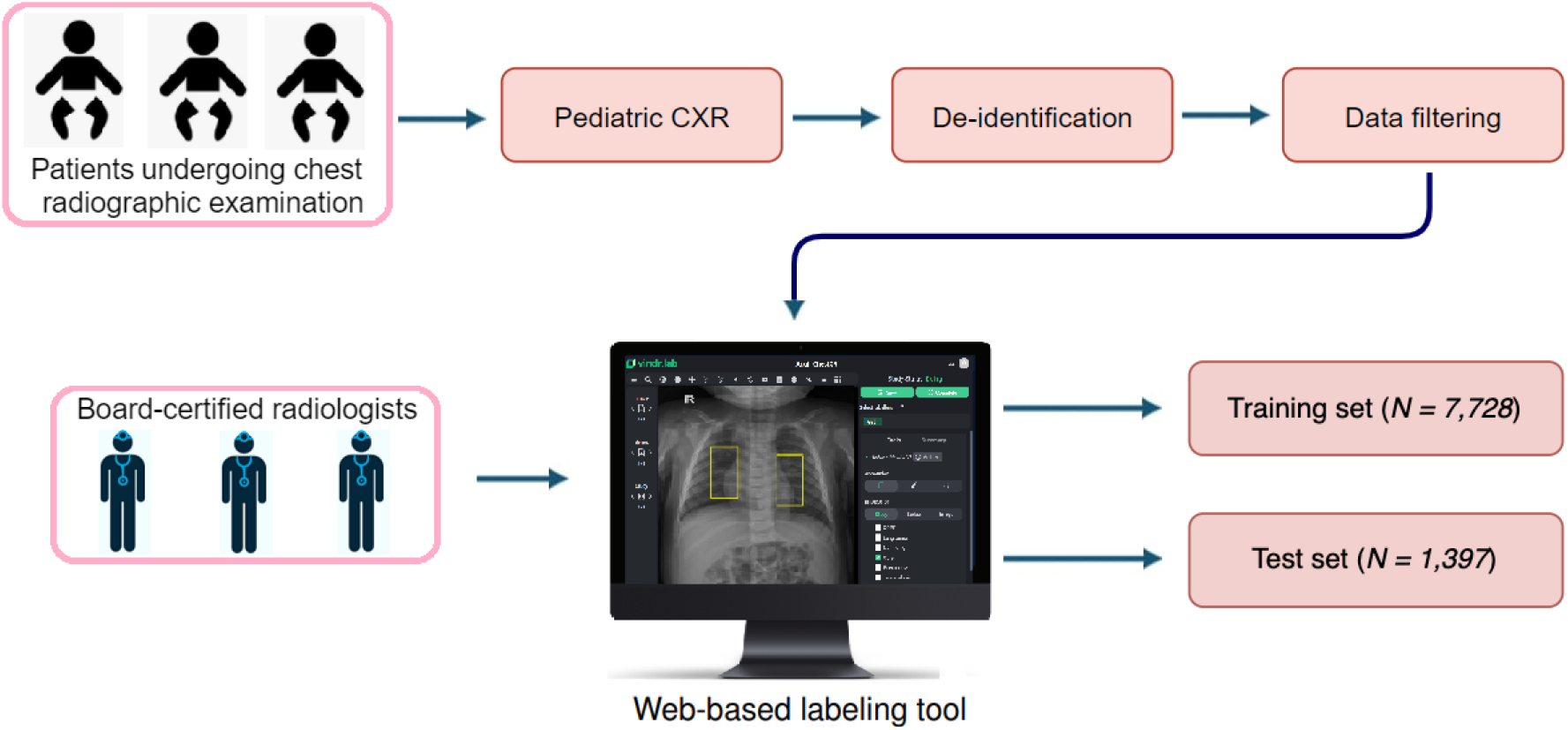
Construction of the VinDr-PCXR dataset: (1) raw pediatric scans in DICOM format were collected retrospectively from the hospital’s PACS at PTOPH. These images were de-identified to protect patient’s privacy; (2) invalid files (including adult CXR images, images of other modalities or other body parts, images with low quality, or incorrect orientation) were manually filtered out; (3) A web-based DICOM labeling tool called VinDr Lab was developed to remotely annotate DICOM data. The annotated dataset was then divided into a training set (*N = 7,728*) and a test set (*N = 1,397*) for algorithm development.

### Data de-identification

In this study, we follow the HIPAA Privacy Rule^20^ to protect most individually identifiable health information from the DICOM images. To this end, we removed or replaced with random values all personally identifiable information associated with the images via a two-stage de-identification process. At the first stage, a Python script was used to remove all DICOM tags of protected health information (PHI)^21^ such as patient’s name, patient’s date of birth, patient ID, or acquisition time and date, etc. For the purpose of loading and processing DICOM files, we only retained a limited number of DICOM attributes that are necessary, as indicated in Table 3 (Supplementary materials). In the second stage, we manually removed all textual information appearing on the image data, i.e., pixel annotations that could include patient’s identifiable information.

### Data filtering

The collected raw data included a significant amount of outliers including CXRs of adult patients, body parts other than chest (abdominal, spine, and others), low-quality images, or lateral CXRs. To filter a large number of CXR scans, we trained a lightweight convolutional neural network (CNN)^22^ to remove all outliers automatically. Next, a manual verification was performed to ensure all outliers had been fully removed.

### Data labeling

The VinDr-PCXR dataset was labeled for a total of 36 findings and 15 diagnoses. These labels were divided into two categories: local labels (#1 – #36) and global labels (#37 – #52). The local labels should be marked with bounding boxes that localize the findings, while the global labels should reflect the diagnostic impression of the radiologist. This list of labels was suggested by a committee of the most experienced pediatric radiologists. To select these labels, the committee took into account two key factors. First, findings and diseases are prevalent. Second, they can be differentiated on pediatric chest X-ray scans. Figure 2 illustrates several samples with both local and global labels annotated by our radiologists.

**Figure 2.**
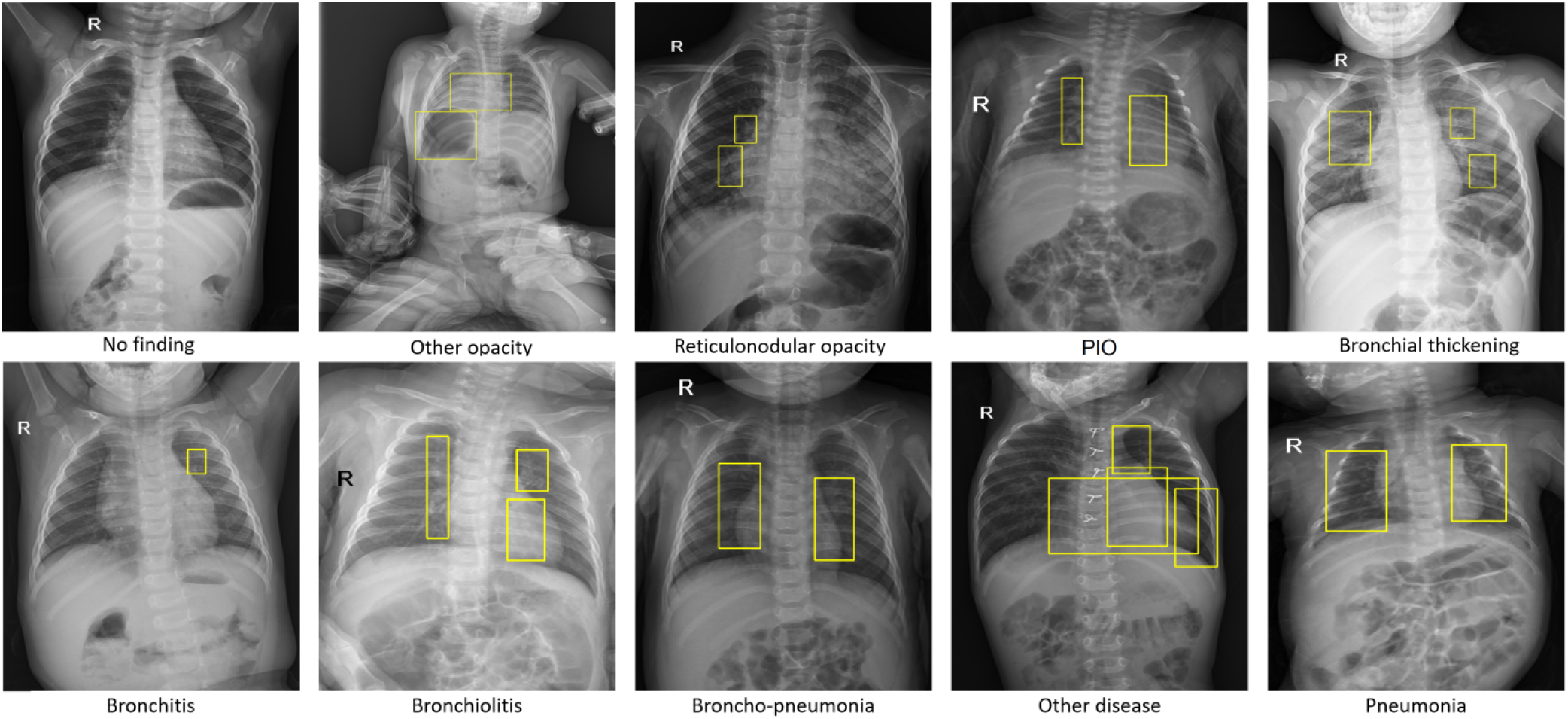
Several examples of pediatric CXR images with radiologist’s annotations. Local labels marked by radiologists are plotted on the original images for visualization purposes. These annotations show abnormal findings from the scans. The global labels, that classify images into diseases, are in bold and listed at the bottom of each example.

To facilitate the labeling process, we designed and built a web-based framework called VinDr Lab (https://vindr.ai/vindr-lab) that allows a team of 3 experienced radiologists remotely annotate the data. All the radiologists participating in the labeling process were certified in diagnostic radiology and received healthcare profession certificates. Each sample in the training set was assigned to one radiologist for annotating. Additionally, all of the participating radiologists were blinded to relevant clinical information. A set of 9,125 pediatric CXRs were randomly annotated from the filtered data, of which 7,728 scans serve as the training set and the remaining 1,397 form the test set.

Once the labeling was completed, the annotations of all pediatric CXRs were exported in JavaScript Object Notation (JSON) format. We developed a Python script to parse JSON files and organized the annotations in the form of a single comma-separated values (CSV) file. Each CSV file contains labels, bounding box coordinates, and their corresponding image identifiers (IDs). The data characteristics, including patient demographic and the prevalence of each finding or disease, are summarized in Table 2. The distributions of abnormal findings and pathologies in the training set are drawn in Figure 3 and Figure 4, respectively.

**Table 2.** Dataset characteristics of VinDr-PCXR.

|  | Characteristics | Training set | Test set |
| --- | --- | --- | --- |
| Collection statistics | Years | 2020 to 2021 | 2020 to 2021 |
|  | Number of scans | 7,728 | 1,397 |
|  | Number of human annotators per scan | 1 | 1 |
|  | Image size (pixel×pixel, median) | 1,643 × 1,349 | 1,638 × 1,343 |
|  | Age (years, median)* | 1.71 | 1.69 |
|  | Male (%)* | 57.63 | 59.14 |
|  | Female (%)* | 42.37 | 40.86 |
|  | Data size (GB) | 30.9 | 5.7 |
| Local labels | 1. Boot-shaped heart (%) | 35 (0.45%) | 6 (0.43%) |
|  | 2. Peribronchovascular interstitial opacity or PIO (%) | 1,358 (17.57%) | 248 (17.75%) |
|  | 3. Reticulonodular opacity (%) | 509 (6.59%) | 90 (6.44%) |
|  | 4. Bronchial thickening (%) | 562 (7.27%) | 116 (8.30%) |
|  | 5. Enlarged PA (%) | 61 (0.79%) | 11 (0.79%) |
|  | 6. Cardiomegaly (%) | 161 (2.08%) | 29 (2.08%) |
|  | 7. Other opacity (%) | 148 (1.92%) | 27 (1.93%) |
|  | 8. Intrathoracic digestive structure (%) | 2 (0.03%) | 0 (0.00%) |
|  | 9. Diffuse aveolar opacity (%) | 119 (1.54%) | 21 (1.50%) |
|  | 10. Other lesion (%) | 65 (0.84%) | 11 (0.79%) |
|  | 11. Consolidation (%) | 176 (2.28%) | 35 (2.51%) |
|  | 12. Mediastinal shift (%) | 5 (0.06%) | 0 (0.00%) |
|  | 13. Anterior mediastinal mass (%) | 5 (0.06%) | 1 (0.07%) |
|  | 14. Other nodule/mass (%) | 10 (0.13%) | 2 (0.14%) |
|  | 15. Dextro cardia (%) | 16 (0.21%) | 3 (0.21%) |
|  | 16. Aortic enlargement (%) | 2 (0.03%) | 0 (0.00%) |
|  | 17. Pleural effusion (%) | 14 (0.18%) | 3 (0.21%) |
|  | 18. Stomach on the right side (%) | 5 (0.06%) | 1 (0.07%) |
|  | 19. Atelectasis (%) | 23 (0.30%) | 3 (0.21%) |
|  | 20. Calcification (%) | 1 (0.01%) | 0 (0.00%) |
|  | 21. Interstitial lung disease - ILD (%) | 14 (0.18%) | 2 (0.14%) |
|  | 22. Lung hyperinflation (%) | 108 (1.40%) | 21 (1.50%) |
|  | 23. Egg on string sign (%) | 12 (0.16%) | 2 (0.14%) |
|  | 24. Pulmonary fibrosis (%) | 1 (0.01%) | 0 (0.00%) |
|  | 25. Infiltration (%) | 11 (0.14%) | 2 (0.14%) |
|  | 26. Lung cavity (%) | 5 (0.06%) | 1 (0.07%) |
|  | 27. Pneumothorax (%) | 4 (0.05%) | 0 (0.00%) |
|  | 28. Edema (%) | 1 (0.01%) | 0 (0.00%) |
|  | 29. Pleural thickening (%) | 2 (0.03%) | 0 (0.00%) |
|  | 30. Clavicle fracture (%) | 5 (0.06%) | 1 (0.07%) |
|  | 31. Chest wall mass (%) | 3 (0.04%) | 0 (0.00%) |
|  | 32. Lung cyst (%) | 8 (0.10%) | 2 (0.14%) |
|  | 33. Emphysema (%) | 1 (0.01%) | 0 (0.00%) |
|  | 34. Bronchiectasis (%) | 3 (0.04%) | 0 (0.00%) |
|  | 35. Expanded edges of the anterior ribs (%) | 2 (0.03%) | 0 (0.00%) |
|  | 36. Paravertebral mass (%) | 2 (0.03%) | 0 (0.00%) |
| Global labels | 37. No finding (%) | 5,143 (66.55%) | 907 (64.92%) |
|  | 38. Bronchitis (%) | 842 (10.90%) | 174 (12.46%) |
|  | 40. Brocho-pneumonia (%) | 545 (7.05%) | 84 (6.01%) |
|  | 41. Other diseases (%) | 412 (5.33%) | 77 (5.51%) |
|  | 42. Bronchiolitis (%) | 497 (6.43%) | 90 (6.44%) |
|  | 43. Situs inversus (%) | 11 (0.14%) | 2 (0.14%) |
|  | 44. Pneumonia (%) | 392 (5.07%) | 89 (6.37%) |
|  | 45. Pleuro-pneumonia (%) | 6 (0.08%) | 0 (0.00%) |
|  | 46. Diaphragmatic hernia (%) | 3 (0.04%) | 0 (0.00%) |
|  | 47. Tuberculosis (%) | 14 (0.18%) | 1 (0.07%) |
|  | 48. Congenital emphysema (%) | 2 (0.03%) | 0 (0.00%) |
|  | 49. CPAM (%) | 5 (0.06%) | 1 (0.07%) |
|  | 50. Hyaline membrane disease (%) | 19 (0.25%) | 3 (0.21%) |
|  | 51. Mediastinal tumor (%) | 8 (0.10%) | 1 (0.07%) |
|  | 52. Lung tumor (%) | 5 (0.06%) | 0 (0.00%) |

**Table 3.** The list of DICOM tags that were retained for loading and processing raw images. All other tags were removed for protecting patient privacy. Details about all these tags can be found from DICOM Standard Browser at https://dicom.innolitics.com/ciods.

| DICOM Tag | Attribute Name | Description |
| --- | --- | --- |
| (0010, 0040) | Patient's Sex | Sex of the named patient. |
| (0010, 1010) | Patient's Age | Age of the patient. |
| (0010, 1020) | Patient's Size | Length or size of the patient, in meters. |
| (0010, 1030) | Patient's Weight | Weight of the patient, in kilograms. |
| (0028, 0010) | Rows | Number of rows in the image. |
| (0028, 0011) | Columns | Number of columns in the image. |
| (0028, 0030) | Pixel Spacing | Physical distance in the patient between the center of each pixel, specified by a numeric pair - adjacent row spacing (delimiter) adjacent column spacing in mm. |
| (0028, 0034) | Pixel Aspect Ratio | Ratio of the vertical size and horizontal size of the pixels in the image specified by a pair of integer values where the first value is the vertical pixel size, and the second value is the horizontal pixel size. |
| (0028, 0100) | Bits Allocated | Number of bits allocated for each pixel sample. Each sample shall have the same number of bits allocated. |
| (0028, 0101) | Bits Stored | Number of bits stored for each pixel sample. Each sample shall have the same number of bits stored. |
| (0028, 0102) | High Bit | Most significant bit for pixel sample data. Each sample shall have the same high bit. |
| (0028, 0103) | Pixel Representation | Data representation of the pixel samples. Each sample shall have the same pixel representation. |
| (0028, 0106) | Smallest Image Pixel Value | The minimum actual pixel value encountered in this image. |
| (0028, 0107) | Largest Image Pixel Value | The maximum actual pixel value encountered in this image. |
| (0028, 1050) | Window Center | Window center for display. |
| (0028, 1051) | Window Width | Window width for display. |
| (0028, 1052) | Rescale Intercept | The value b in relationship between stored values (SV) and the output units specified in Rescale Type (0028,1054). Each output unit is equal to $m \cdot SV + b$. |
| (0028, 1053) | Rescale Slope | Value of m in the equation specified by Rescale Intercept (0028,1052). |
| (7FE0, 0010) | Pixel Data | A data stream of the pixel samples that comprise the image. |
| (0028, 0004) | Photometric Interpretation | Specifies the intended interpretation of the pixel data. |
| (0028, 2110) | Lossy Image Compression | Specifies whether an image has undergone lossy compression (at a point in its lifetime). |
| (0028, 2114) | Lossy Image Compression Method | A label for the lossy compression method(s) that have been applied to this image. |
| (0028, 2112) | Image Compression Ratio | Describes the approximate lossy compression ratio(s) that have been applied to this image. |
| (0028, 0002) | Samples per Pixel | Number of samples (planes) in this image. |
| (0028, 0008) | Number of Frames | Number of frames in a multi-frame image. |

**Figure 3.**
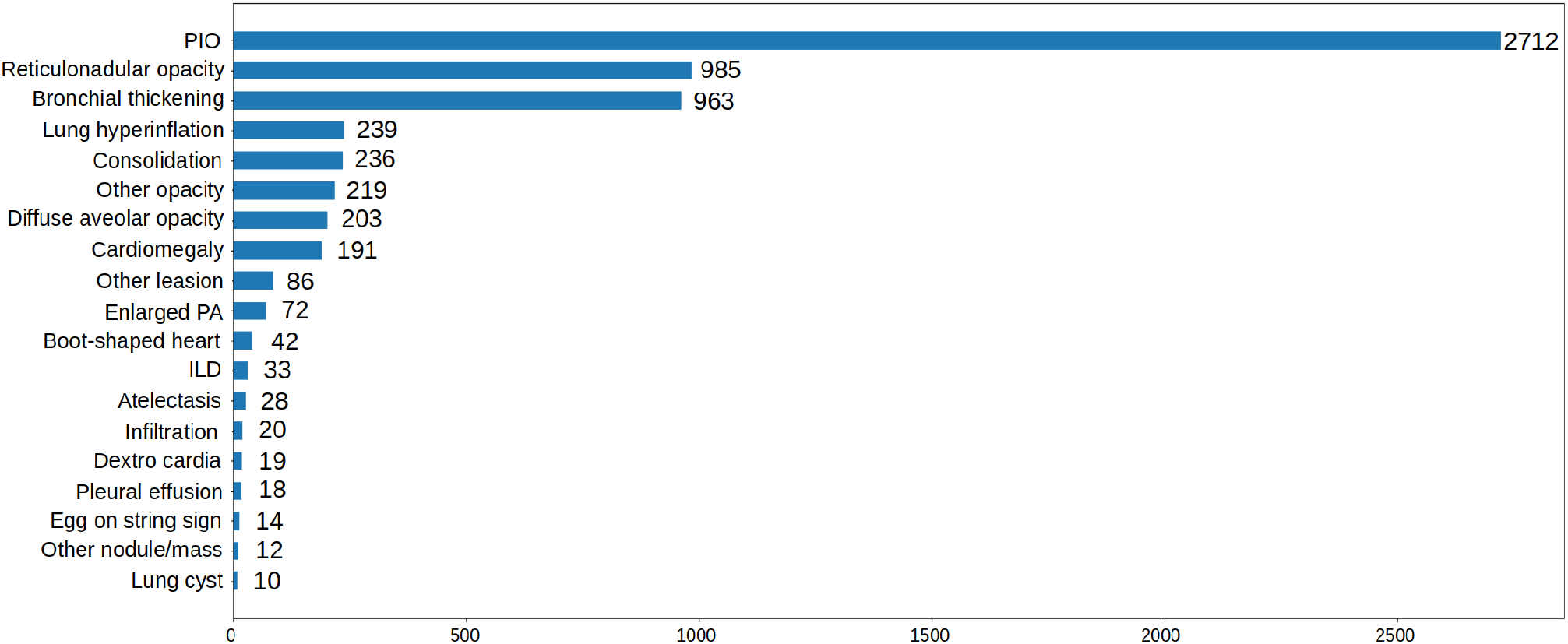
Distribution of abnormal findings on the training set of VinDr-PCXR. Rare findings (less than 10 examples) are not included.

**Figure 4.**
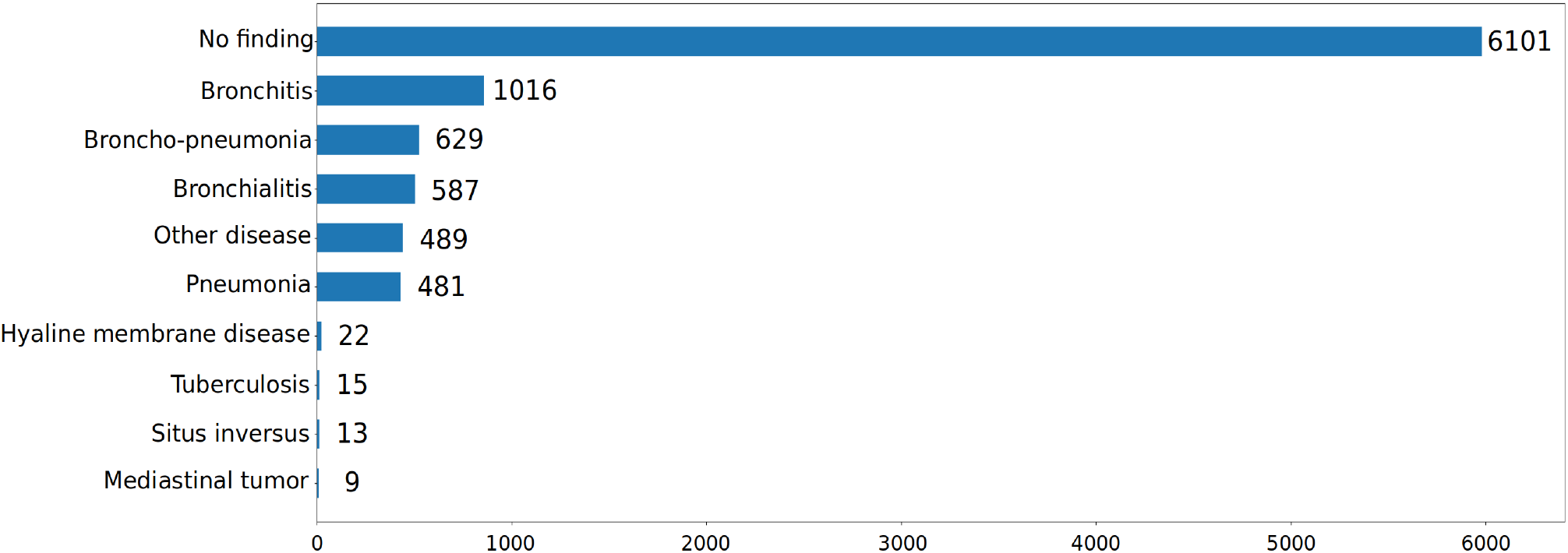
Distribution of pathologies on the training set of VinDr-PCXR. Rare diseases (less than 10 examples) are not included.

## Data Records

The VinDr-PCXR dataset will be made available for public download on PhysioNet (https://physionet.org/). We offer complete imaging data as well as ground truth labels for both the training and test datasets. The pediatric scans were split into two folders: one for training and one for testing, named as “`train`” and “`test`”, respectively. The value of the SOP Instance UID provided by the DICOM tag (0008, 0018) was encoded into a unique, anonymous identifier for each image. To this end, we used the Python `hashlib` module (see Code Availability) to encode the SOP Instance UIDs into image IDs. The radiologists’ local annotations of the training set were provided in a CSV file called `annotations_train.csv`. Each row of the CSV file represents a bounding box annotation with the following attributes: image ID (`image_id`), radiologist ID (`rad_id`), label’s name (`class_name`), bounding box coordinates (`x_min`, `y_min`, `x_max`, `y_max`), and label class ID (`class_id`). Here, the coordinates of the box’s upper-left corner are (`x_min`, `y_min`), and the coordinates of the box’s lower right corner are (`x_max`, `y_max`). Meanwhile, the image-level labels of the training set were stored in a different CSV file called `image_labels_train.csv`, with the following fields: Image ID (`image_id`), radiologist ID (`rad_ID`), and labels (`labels`) for both the findings and diagnoses. Each image ID is associated with a vector of multiple labels corresponding to different pathologies, with positive pathologies encoded as “`1`” and negative pathologies encoded as “`0`”. Similarly, the test set’s bounding-box annotations and image-level labels were saved in the files `annotations_test.csv` and `image_labels_test.csv`, respectively.

## Technical Validation

The data de-identification process was controlled. Specifically, all DICOM meta-data was parsed and manually reviewed to ensure that all individually identifiable health information (PHI)^21^ of the children patients has been removed to meet the U.S. HIPAA^20^ regulations. In addition, pixel values of all pediatric CXR scans were also carefully examined by human readers. During this review process, all scans were manually reviewed case-by-case by a team of 10 human readers. A small number of images containing private textual information that had not been removed by our algorithm was excluded from the dataset. The manual review process also helped identify and discard out-of-distribution samples such as CXRs of adult patients, body parts other than the chest, low-quality images, or lateral CXRs that our machine learning classifier was not able to detect. A set of rules underlying our web-based annotation tool were developed to control the quality of the labeling process. These rules prevent human annotators from mechanical mistakes like forgetting to choose global labels or marking lesions on the image while choosing “`No finding`” as the global label.

## Usage Notes

The VinDr-PCXR dataset was established for the purpose of developing and evaluating machine learning algorithms for detecting and localizing anomalies in pediatric CXR images. The dataset has been previously used in a study on the diagnosis of multiple diseases in pediatric patients^23^ and showed promising results. The primary uses for which the VinDr-PCXR dataset was conceptualized include:

- Developing and validating a predictive model for the classification of common thoracic diseases in pediatric patients.
- Developing and validating a predictive model for the localization of multiple abnormal findings on the pediatric chest X-ray scans.

Finally, the released dataset remains with limitations that still need to be addressed in the future, including:

- The dataset did not contain clinical information associated with DICOM images, which is essential for the interpretation of CXR in children patients.
- The number of examples for rare diseases (e.g., Congenital pulmonary airway malformation (CPAM), Congenital emphysema, Diagphramatic hernia, Mediastinal tumor, Pleuro-pneumonia, Situs inversus, Lung tumor) or findings (Emphysema, Edema, Calcification, Chest wall mass, Bronchectasis, Pleural thickening, Clavicle fracture, Pleuropulmonary mass, Paraveterbral mass, etc.) are limited. Hence, training supervised learning algorithms, which requires a large-scale annotated dataset, on the VinDr-PCXR dataset to diagnose the rare diseases and findings is not reliable.

To download and use the VinDr-PCXR, users are required to accept the PhysioNet Credentialed Health Data License 1.5.0. By accepting this license, users agree that they will not share access to the dataset with anyone else. For any publication that explores this resource, the authors must cite this original paper and release their code and models.

## Data Availability

All data produced are available online at https://physionet.org/

## Code Availability

This study used the following open-source repositories to load and process DICOM scans: Python 3.7.0 (https://www.python.org/); Pydicom 1.2.0 (https://pydicom.github.io/); OpenCV-Python 4.2.0.34 (https://pypi.org/project/opencv-python/); and Python hashlib (https://docs.python.org/3/library/hashlib.html). The code for data de-identification was made publicly available at https://github.com/vinbigdata-medical/vindr-cxr. The code to train CNN classifier for the out-of-distribution task was made publicly available at https://github.com/vinbigdata-medical/DICOM-Imaging-Router.

## Acknowledgements

The collection of this dataset was funded by the Smart Health Center, VinBigData JSC. The authors would like to acknowledge the Phu Tho Obstetric & Pediatric Hospital for agreeing to make the VinDr-PCXR dataset publicly available. We are especially thankful to Anh T. Nguyen, Huong T.T. Nguyen, Ngan T.T. Nguyen for their helps in the data collection and labeling process.

## Author contributions

H.Q.N. and H.H.P designed the study; T.T.T. performed the data de-identification; H.Q.N., and H.H.P. wrote the paper; all authors reviewed the manuscript.

## Supplementary materials

